# Limited Validity of Mayo Endoscopic Subscore in Ulcerative Colitis with Concomitant Primary Sclerosing Cholangitis

**DOI:** 10.1101/2024.04.08.24305005

**Authors:** Pavel Wohl, Alzbeta Krausova, Petr Wohl, Ondrej Fabian, Lukas Bajer, Jan Brezina, Pavel Drastich, Mojmír Hlavaty, Petra Novotna, Michal Kahle, Julius Spicak, Martin Gregor

**Affiliations:** Department of Gastroenterology, Institute for Clinical and Experimental Medicine, Prague, Czech Republic; Laboratory of Integrative Biology, Institute of Molecular Genetics of the Czech Academy of Sciences, Prague, Czech Republic; Diabetes Center, Institute for Clinical and Experimental Medicine, Prague, Czech Republic; Department of Clinical and Transplant Pathology, Institute for Clinical and Experimental Medicine, Prague, Czech Republic; Department of Pathology and Molecular Medicine, 3rd Faculty of Medicine, Charles University and Thomayer Hospital, Prague, Czech Republic; Department of Data Analysis, Statistics, and AI, Institute for Clinical and Experimental Medicine, Prague, Czech Republic

**Keywords:** Primary sclerosing cholangitis with ulcerative colitis, diagnosis, Nancy histological index, Mayo endoscopic subscore

## Abstract

**Background and Aims:** Ulcerative colitis (UC) with concomitant primary sclerosing cholangitis (PSC) represents a distinct disease entity (PSC-UC). Mayo endoscopic subscore (MES) is a standard tool for assessing disease activity in UC but its relevance in PSC-UC remains unclear. Here, we sought to compare MES in a cohort of UC and PSC-UC patients and assess the accuracy using histological activity scoring (Nancy histological index; NHI).

**Methods:** MES was assessed in 30 PSC-UC and 29 UC adult patients during endoscopy. NHI and inflammation were evaluated in biopsies from the caecum, rectum, and terminal ileum. In addition, perinuclear anti-neutrophil cytoplasmic antibodies, fecal calprotectin, body mass index, and other relevant clinical characteristics were collected.

**Results:** The median MES and NHI were similar for UC patients (MES grade 2 and NHI grade 2 in the rectum), but were different for PSC-UC patients (MES grade 0 and NHI grade 2 in the caecum). There was a correlation between MES and NHI for UC patients (Spearman’s ρ = 0.40, p = 0.029), but not for PSC-UC patients. Histopathological examination revealed persistent microscopic inflammation in 88% of PSC-UC patients with MES grade 0 (46% of all PSC-UC patients). Moreover, MES overestimated the severity of active inflammation in another 11% of PSC-UC patients.

**Conclusion:** MES fails to identify microscopic signs of inflammation in the context of PSC-UC. This indicates that histological evaluation should become an integral part of the diagnostic and grading system in both PSC-UC and PSC.

## 1. Introduction

Primary sclerosing cholangitis (PSC) is a chronic cholestatic liver disease characterized by progressive inflammation, fibrosis, and diffuse multiple stricturing of the intrahepatic and extrahepatic bile ducts^1^. In up to 80% of patients, PSC is closely associated with inflammatory bowel disease (IBD), prevalently with a unique type of ulcerative colitis (UC) known as PSC-UC^2, 3^.

As a distinct clinical phenotype, PSC-UC manifests with colonoscopic features that differ from those of typical UC without hepatobiliary disease. Interestingly, PSC-UC may not develop clinically apparent gastrointestinal symptoms^4, 5^. Multiple studies^2, 6, 7^ have shown that the colonic inflammation in PSC-UC is typically more pronounced in the right-sided colon with often minimal to normal mucosal findings in the rectum. Furthermore, PSC-UC is characterized by a lower incidence of inflammatory polyps^4, 8^ and a higher incidence of backwash ileitis^9^ when compared to UC. In up to 94% of PSC-UC cases, the phenotype is reported as pancolitis with rectal sparing. Although colitis in PSC tends to follow a quiescent course, PSC-UC is associated with a high incidence of malignancies represented mainly by colitis-associated carcinoma (CAC)^10–13^. The risk of CAC is higher in PSC-UC than in UC alone, which is why accurate diagnosis is important for all PSC-UC patients.

IBD diagnosis is largely based on clinical symptoms, endoscopy, and histopathology^14^ with endoscopic assessment being the most feasible and reliable approach^15^ in routine clinical practice. Among many different endoscopic scores for UC^16^, the Mayo Endoscopic Subscore (MES)^17, 18^, a component of the Mayo Clinic Score, is recommended to assess disease activity and remains the most frequently used score in both clinical practice and clinical trials^17^. Surprisingly, despite accumulated evidence of limited diagnostic accuracy of endoscopic techniques^19–23^, namely in the context of mild mucosal inflammation, neither MES nor any of the other endoscopic scores have been validated for PSC-UC.

Endoscopy based approaches are not sufficiently reliable for PSC-UC patients. This can lead to poor therapeutic decisions and misguided treatment. Histopathological evaluation can remedy this by detecting potential microscopic disease activity, despite the absence of clinical or endoscopic signs of disease common in PSC-UC patients^19–23^. Out of the more than 30 described UC histological scores^24^, the newly established Nancy Histological Index (NHI)^24–26^, has quickly become one of the most popular histological scoring systems of inflammatory activity in UC. In 2020, the European Crohn’s and Colitis Organization recommended NHI for daily clinical practice^27^.

Given challenges of endoscopy for PSC-UC, the diagnostic relevance of MES for PSC-UC is unclear, despite MES being a standard tool for assessing inflammation in UC. The overall objective of this study was 1) to compare the reliability of MES as a diagnostic tool between UC and PSC-UC patient cohorts and 2) to assess the accuracy of MES in PSC-UC patients using histological disease activity scoring (NHI).

## 2. Methods

### 2.1 Patients

This study was a prospective longitudinal performed at the Institute for Clinical and Experimental Medicine (Prague, Czech Republic), a tertiary health care center. We included 59 Caucasian adult patients diagnosed with UC (n=29) and PSC-UC (n=30) according to conventional diagnostic criteria, who were admitted to the Hepatogastroenterology Department for a colonoscopy from July 2016 to March 2021. As portal hypertension with portal colopathy in liver cirrhosis are common endoscopic features that may mimic some inflammatory changes typical for PSC-UC, patients with advanced liver cirrhosis with portal hypertension were excluded. Other exclusion criteria were colitis-associated cancer and colonic dysplasia. Study cohort consisted of 20 females and 39 males (sex). This study was approved by the Ethics Committee of the Institute for Clinical and Experimental Medicine and Thomayer Hospital with Multi-Center Competence (G16-06-25) and performed in accordance with the Declaration of Helsinki. Written informed consents were obtained from all subjects before the study. All patients have been characterized as summarized in Table 1.

**Table 1.** Characterization of patients.

|  | UC | PSC-UC | p |
| --- | --- | --- | --- |
| Patients, *n* | 29 | 30 | - |
| Sex (male), *n* (%) | 16 (55) | 23 (77) | 0.10 |
| Age (years), mean ± SD | 43.2 ± 12.5 | 37.2 ± 2.9 | 0.07 |
| Duration of disease (years), mean ± SD | 12.9 ± 10.6 | 10.6 ± 6.9 | 0.62 |
| pANCA positive, *n* (%) | 14 (48) | 16 (53) | 0.8 |
| Portal hypertension, *n* | 0 | 0 | - |
| Cholecystectomy, *n* (%) | 1 (3) | 1 (3) | - |
| BMI (a. u.), mean ± SD | 25.1 ± 4.1 | 23.5 ± 4.2 | 0.1 |
| Corticosteroids, *n* (%) | 10 (34) | 10 (33) | - |
| Mesalamine, *n* (%) | 12 (41) | 20 (66) | - |
| Mesalamine and immunomodulators, *n* (%) | 12 (41) | 9 (30) | - |
| Biological treatment, *n* (%) | 5 (17) | 0 | - |
| Ursodeoxycholic acid, *n* (%) | 0 | 30 (100) | - |
| Fecal calprotectin (μg/g), median (95% CI) | 478 (263–917) | 96 (60-231) | <0.001 |

UC, ulcerative colitis; PSC-UC, primary sclerosing cholangitis with concomitant ulcerative colitis; pANCA, perinuclear anti-neutrophil cytoplasm autoantibodies; BMI, body mass index; a.u., arbitrary units; SD, standard deviation; CI, confidence interval

### 2.2 Endoscopic and histological evaluations

All UC and PSC-UC patients were subjected to a colonoscopy with a standard white light endoscope. During the colonoscopy, two or three biopsies from the terminal ileum, caecum, and rectum of endoscopically most severely inflamed mucosa were collected. Endoscopic disease activity was assessed using MES^17^. Histological inflammation in the colon biopsies (caecum and rectum) was assessed by NHI as published previously for UC^25,26^. In addition, histological inflammation in biopsies from the terminal ileum was determined by a four-grade scoring system (0-3), where 0 corresponds to normal and 3 to severe inflammation. Blinded histopathological evaluation of paraffin-embedded sections stained with hematoxylin-eosin was performed by a trained pathologist (O.F.).

### 2.3 Primary sclerosing cholangitis

PSC was defined by the presence of intra- and/or extra-hepatic bile duct abnormalities in the form of beading, duct ectasia, and stricturing of the intra- or extra-hepatic bile ducts documented in the medical record from endoscopic retrograde cholangiopancreatography, magnetic resonance cholangiopancreatography, and/or liver biopsy. Small duct PSC was defined when there were histological features consistent with PSC on liver biopsy in the absence of characteristic radiological features. The diagnosis of PSC was also confirmed by laboratory tests (see below).

### 2.4 Laboratory and biochemical parameters

Blood analysis was performed on the day of the colonoscopy, including the determination of haemoglobin, leucocytes, platelets, and albumin (not shown). A stool sample that was obtained immediately before bowel preparation was provided by each patient for the analysis of faecal calprotectin (FC). FC level was measured by ELISA EliA kit (Phadia AB, Uppsala, Sweden). Detection of anti-neutrophil cytoplasmic antibody (ANCA) and IgG4 was performed using kits from Inova Diagnostics Inc., San Diego, USA.

### 2.5 Statistical analyses

All ordinal variables are presented as medians with 95% confidence intervals; continuous variables are expressed as means ± SEM. All differences between independent ordinal variables were tested by Mann-Whitney U test, differences in paired measurements were assessed by Wilcoxon signed-rank test and differences in proportions were tested by Fisher exact test. Correlations are expressed as Spearman’s rank correlation coefficient (ρ). For comparison of MES with NHI, only the most severely affected lesion with the highest NHI grade was considered for each patient. All data were analyzed using the Python ecosystem. Statistical significance was accepted at p ≤ 0.05.

## 3. Results

### 3.1 MES and NHI scoring

The severity of UC was assessed both endoscopically and histologically in PSC-UC (Figure 1, upper panels) and UC (Figure 1, lower panels) patients. Endoscopy and histopathology involved examination of samples from the caecum, rectum and ileum. Among UC patients, only 2 individuals (7%) showed normal endoscopic findings (MES grade 0); the majority exhibited mild to moderate mucosal inflammation and damage (MES grades 1 and 2; median MES grade 2; Table 2). Similarly, histopathological examination revealed that most UC patients (90%) showed normal findings or mild inflammation with median NHI grade 1 in caecum (Table 2). However, in the rectum, 30% UC patients exhibited a shift towards moderately to severely active inflammation (median NHI grade 2; Table 2), which aligned with the MES findings. When considering the most severely inflamed biopsy for each patient (Figure 1, “caecum and rectum” panel), a significant correlation between MES and NHI was observed in UC patients (Spearman’s ρ=0.40, p=0.029).

**Figure 1.**
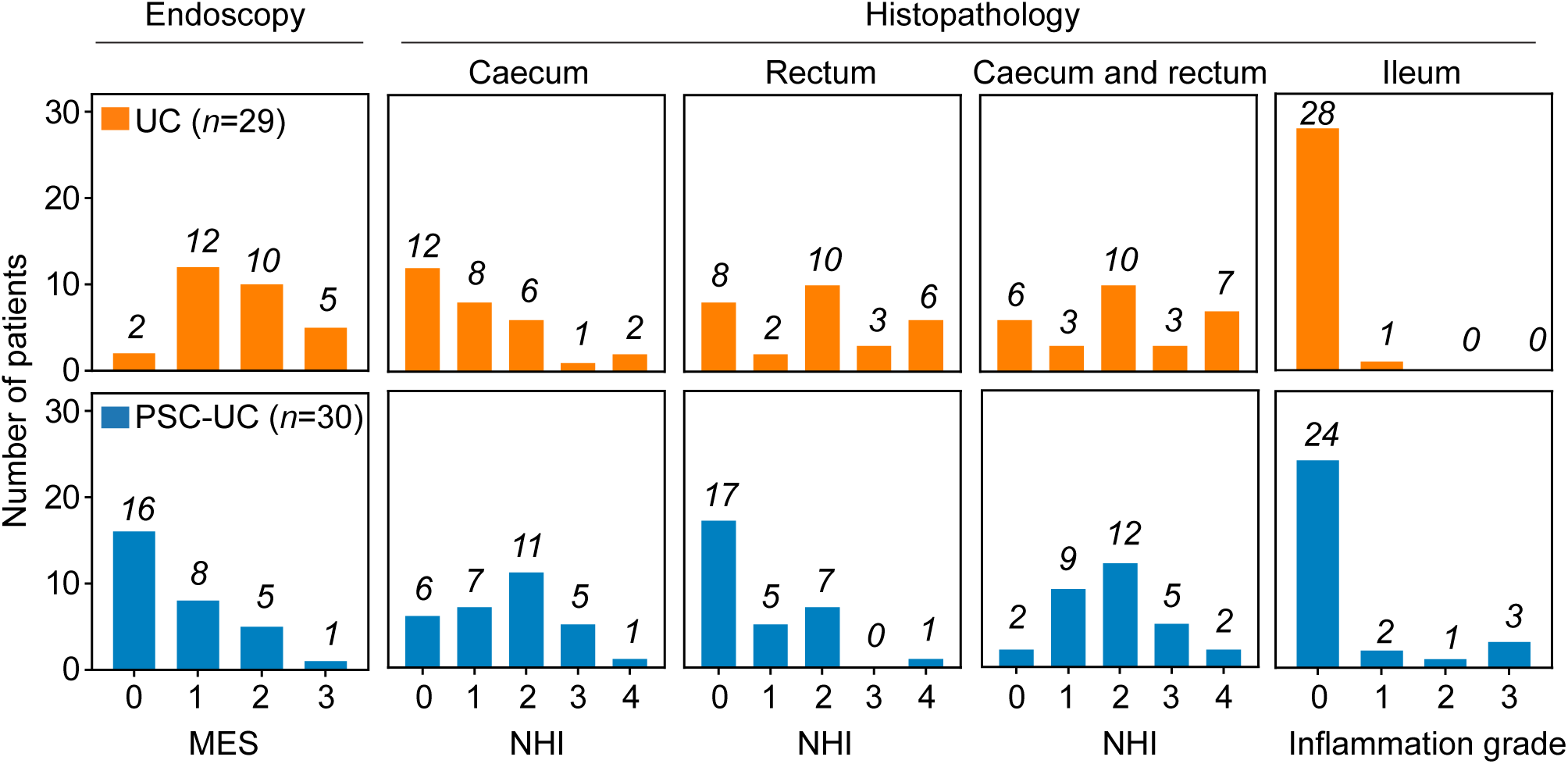
MES, NHI and inflammation grade of PSC-UC and UC patients in the caecum, rectum and ileum. ‘Caecum and rectum’ panels represent only the most severely affected lesions for each patient. The numbers of patients per grade are indicated in the graph.

**Table 2.** Calculated medians (95% CI) of MES and NHI.

|  | PSC-UC | UC | p |
| --- | --- | --- | --- |
| MES | 0 (0-1) | 2 (1-2) | <0.001 |
| inflammation<br>(ileum) | 0 (0-0) | 0 (0-0) | - |
| NHI (caecum) | 2 (1-2) | 1 (0-2) | 0.048 |
| NHI (rectum) | 0 (0-1) | 2 (0-3) | 0.002 |

CI, confidence interval; MES, Mayo endoscopic subscore; NHI, Nancy histological index; PSC-UC, primary sclerosing cholangitis with concomitant ulcerative colitis; UC, ulcerative colitis

Contrary to only 2 UC patients, MES identified 16 (53%) PSC-UC patients with normal endoscopic appearance (MES grade 0; median MES grade 0; Table 2). In the rectum, a similar distribution was observed, with a majority (57%) of PSC-UC patients without histologically active disease (median NHI grade 0, Table 2). However, in the caecum, a more pronounced inflammatory pattern emerged, with 80% of PSC-UC patients displaying NHI grades ranging from 1 to 4 (median NHI grade 2; Table 2). This highlights a significant discrepancy between MES and NHI in PSC-UC patients, which is further corroborated by the lack of correlation between the two measures (Spearman’s ρ=0.27, p=0.14). Notably, for the correlation analysis, only the grades corresponding to the most affected lesions were considered, effectively approximating the situation in the caecum (Figure 1, “caecum and rectum”).

### 3.2 MES and NHI in PSC-UC

To understand the discrepancy between MES and NHI in PSC-UC patients, the NHI values of 16 PSC-UC patients with MES grade 0 were analyzed (Figure 2, upper panels). Surprisingly, this analysis revealed that 13 of these patients (81%) had NHI grades between 1 and 4 in the caecum (Figure 3A,B) and 9 (56%) had NHI grades between 1 and 4 in the rectum. When considering both the caecum and rectum for each patient, only 2 individuals (12.5%) had NHI grades of 0 in both segments (Figure 2, “caecum and rectum” panel). Hence, MES failed to detect active UC in 14 (88%) of the PSC-UC patients with MES grade 0, which corresponds to 46% of all PSC-UC patients.

**Figure 2.**
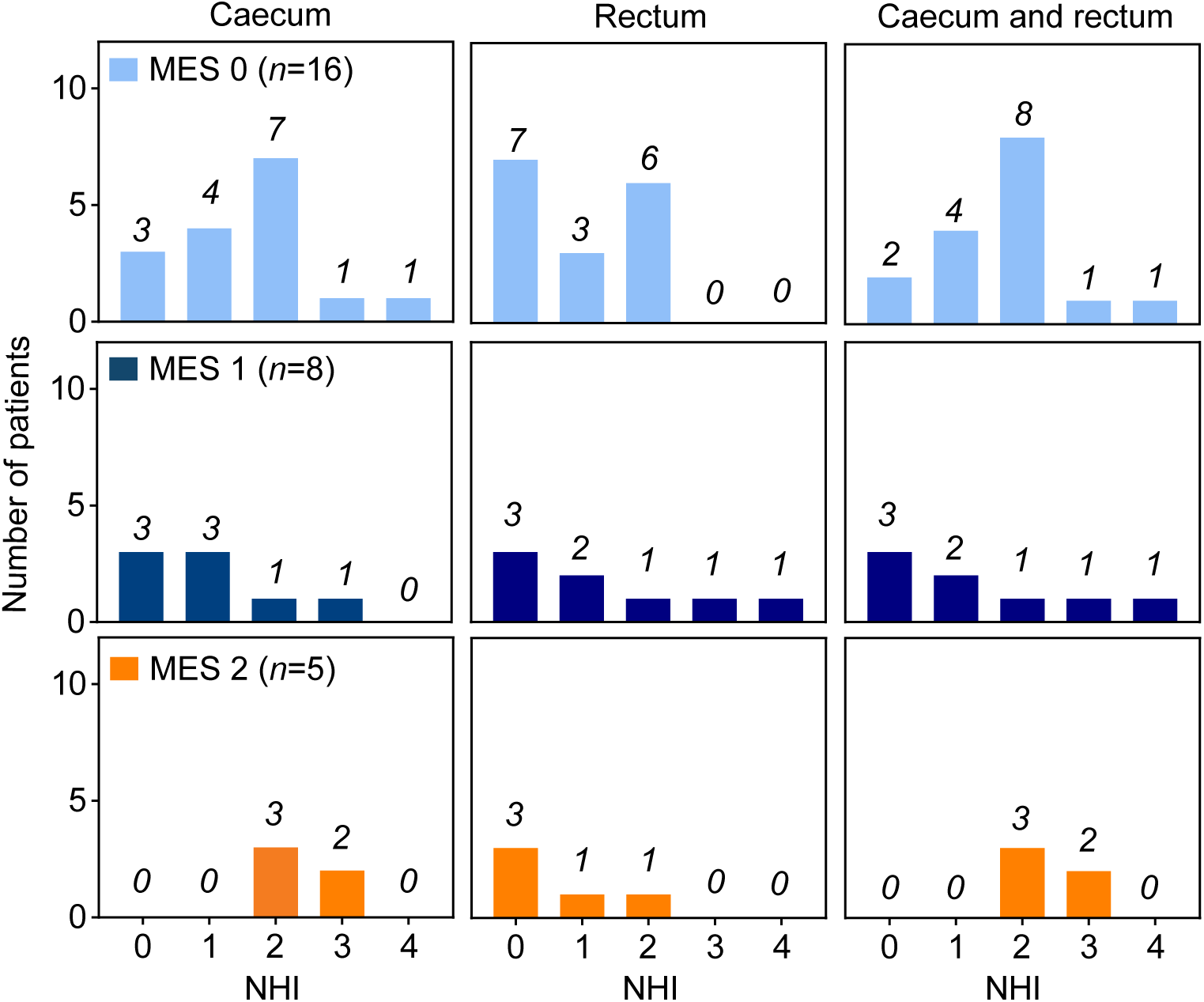
NHI scores of PSC-UC patients with MES grade 0 (top panels), MES grade 1 (middle panels) and MES grade 2 in the caecum and rectum. ‘Caecum and rectum’ panels represent only the most severely affected lesions for each patient. The numbers of patients per grade are indicated in the graph.

**Figure 3.**
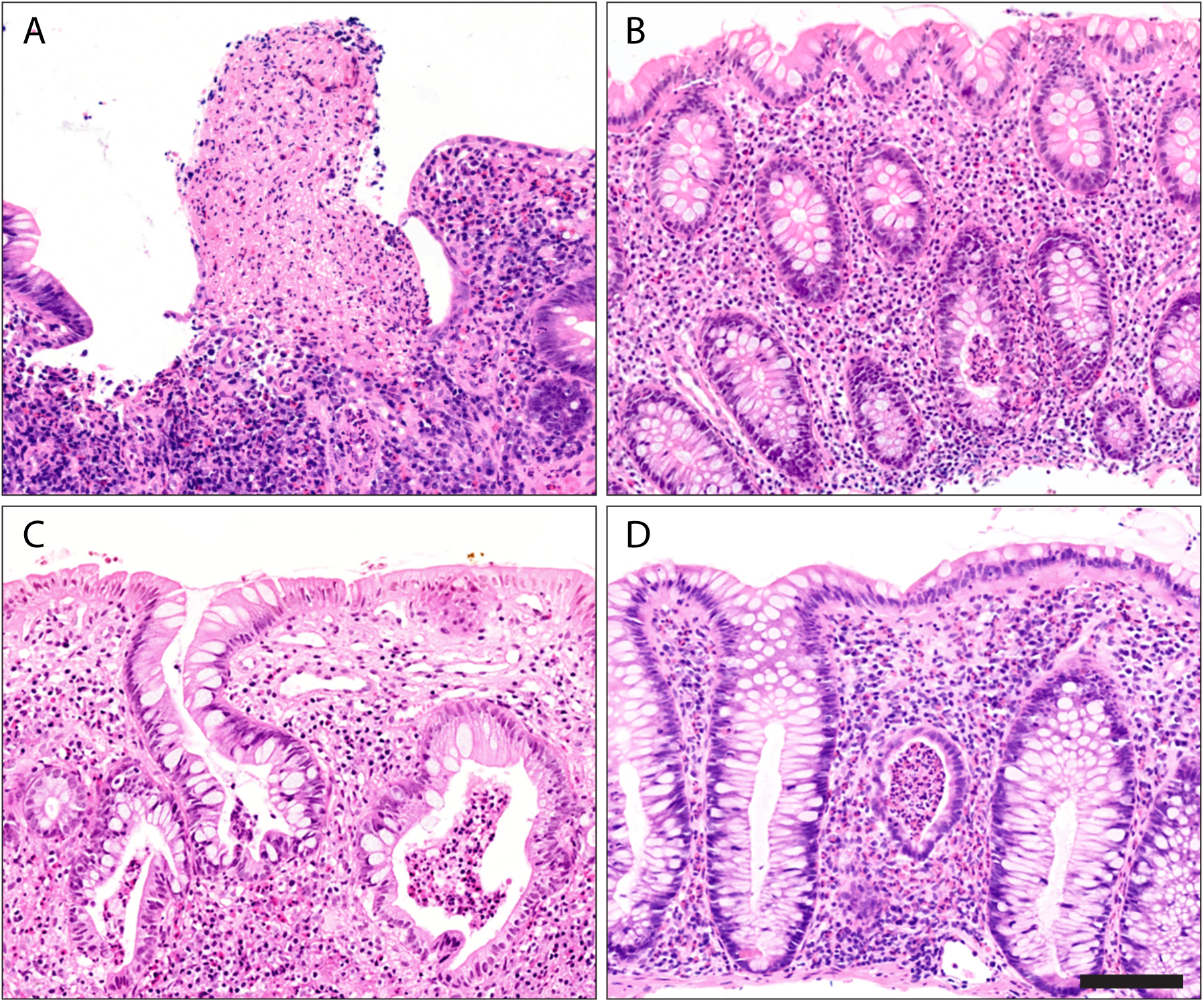
Hematoxylin-eosin stained caecal biopsies from PSC-UC patients with MES grade 0 (A, B), MES grade 1 (C), and MES grade 2 (D). All samples show an active colitic pattern with numerous neutrophils infiltrated into lamina propria and epithelium accompanied by ulceration (A) and crypt abscesses (B-D). Samples were thus assigned NHI grades 4 (A) and 3 (B-D). Scale bar, 50 μm.

Further analysis of the NHI scores revealed that among the 8 PSC-UC patients with MES grade 1, 3 patients showed no signs of inflammation (NHI grade 0) while 3 exhibited active inflammation with NHI grades ranging from 2 to 4 (Figure 2, middle panels; Figure 3C). Similarly, among the 5 patients with MES grade 2, 2 individuals displayed active inflammation with NHI grade 3 (Figure 2, lower panels; Figure 3D). This suggests that MES incorrectly diagnosed another 3 PSC-UC patients. Taken together, these results clearly indicate that MES insufficiently identified ongoing microscopic inflammation in 17 (57%) of PSC-UC patients, which would negatively affect therapeutic decisions.

### 3.3 Backwash ileitis and rectal sparing in PSC-UC patients

All patients were characterized as summarized in Table 1. Of note, fecal calprotectin (FC) levels in UC patients were significantly elevated, with a median of 478 μg/g (CI 263–917), compared to the widely accepted upper limit of 100 μg/g^28^. In contrast, FC levels in PSC-UC patients were within the normal range, with a median of 96 μg/g (CI 60-231). This suggests that PSC-UC patients have nearly absent or very mild UC manifestation, compared to the severe manifestation in UC patients.

Histopathological examination consistently supported the notion of a more severe inflammatory pattern in the rectum of UC patients, as evidenced by the highest NHI grades (Figure 1, Table 2). In contrast, PSC-UC patients predominantly exhibited inflammatory changes in the caecum (median NHI grade 2 vs. 0 in rectum; p<0.01; Figure 1; Table 2), suggesting complete or partial rectal sparing. Indeed, 60% of PSC-UC patients had spared rectal mucosa compared to just 10% of UC patients (p<0.001).

In addition, inflammation scoring in the ileum exhibited similar median values in both UC and PSC-UC patients (median NHI grade 0; Figure 1, Table 2). Nevertheless, 6 PSC-UC patients (20%) exhibited mild to severe inflammation (NHI grades 1-3) compared to only 1 patient in the UC group (3%; p=0.047; Figure 1) thus suggesting higher incidence of backwash ileitis in PSC-UC patients. These findings collectively underscore the variability of inflammation intensity across different segments of the intestines in both PSC-UC and UC.

## 4. Discussion

Up to now, almost 30 scoring systems have been introduced to accurately assess the colitic disease severity in inflammatory bowel diseases. However, despite extensive validations, none of these systems have been universally recognized as versatile^29^. In this study, we aimed to validate the commonly used MES system for evaluating UC in PSC-UC patients and compare the results with histopathological observations expressed through the NHI. Our findings demonstrate a correlation between MES and NHI in UC patients, but a lack of correlation in PSC-UC patients (as indicated by Spearman’s ρ). MES fails to identify ongoing histological inflammation (NHI grade 1-4, Figure 2 and 3) in more than 46% of PSC-UC patients who were assigned MES grade 0. This significant discrepancy highlights a major limitation of endoscopic assessment, which could potentially lead to an underestimation of the severity of PSC-UC. Therefore, we recommend the routine use of histological scoring in clinical practice to overcome this limitation.

It has been reported that UC in PSC manifests mildly or even completely without symptoms compared to typical UC^5, 30^. Additionally, Murasugi et al.^5^ documented no correlation between the severity of liver disease and colonic inflammation expressed by MES. They concluded that it is important for colonoscopy to be routinely performed immediately following a diagnosis of PSC. However, our observations show that histopathological evaluation should always accompany colonoscopy to avoid underdiagnosis.

In our current study, the NHI revealed more severe colitis in the caecum of PSC-UC patients, along with an increased occurrence of backwash ileitis and rectal sparing. These findings are consistent with previous studies that have reported a high prevalence of pancolitis in PSC-UC patients, ranging from 80% to 95%, with varying degrees of severity and a pronounced localization in the right-sided colon^4, 5, 7, 30^. Therefore, it is crucial to inspect all colonic segments (ileum, caecum, rectum, sigmoid, descending, transverse and ascending colon) using both colonoscopy and histopathology to obtain a comprehensive understanding of the disease presentation in PSC-UC patients.

In addition, we suggest that the MES should be evaluated individually for each colonic segment. Lobatón et al.^32^, previously developed a modified MES (MMES) that takes into account the extent and severity of endoscopic activity in UC. While the MMES is complex and informative, our experience indicates that it is more suitable for use in clinical trials rather than in everyday clinical practice. Therefore, we propose that MES scores, along with NHI expressed separately for each colonic segment, serve as ideal and informative prognostic factors for assessing complex disease activity.

Although NHI has not been previously validated for PSC-UC, it has been validated for UC^26^. Furthermore, it has shown a good correlation with other established indices such as the Geboes score and global visual analog scale^24^. Given this correlation, we have successfully employed NHI to evaluate the microscopic disease activity in PSC-UC. Importantly, interpretation must be exercised cautiously especially in treated patients, as NHI grade 0 is assigned to both normal histological observation and a mild chronic inflammation without activity.

In conclusion, our study highlights the limited validity of the standard MES scoring system in the context of PSC-UC. Although our study involved a small number of patients from a single center, it emphasizes the importance of incorporating histological evaluation into the diagnostic and grading system for PSC and PSC-UC. Early diagnosis is crucial for the overall clinical management of patients, particularly in terms of monitoring malignant complications and determining the need for orthotopic liver transplantation. Therefore, we recommend that the MES scoring system be reserved for assessing the endoscopic disease activity of UC without PSC, while histological evaluation should be considered an essential component in the UC diagnosis of PSC and PSC-UC patients.

## Acknowledgments

We thank Lenka Bruhova and Katerina Dvorakova for their excellent support. We are especially grateful to all the patients participating in this study.

## Author Contributions

Pa.W. designed the study; T.H., L.B., P.D., Pe.W., M.H. recruited and treated patients, collected and analyzed data; O.F. performed histological evaluations; P.B. analyzed data; M.K. performed statistical analyses; all authors interpreted data; Pa.W., A.K., M.G. wrote the manuscript.

## Data Availability

The data underlying this article cannot be shared publicly due to the privacy of individuals that participated in this study. The data will be shared on reasonable request to the corresponding authors.

## Ethics approval

This study was approved by the Ethics Committee of the Institute for Clinical and Experimental Medicine and Thomayer Hospital with Multi-Center Competence (G16-06-25) and performed in accordance with the Declaration of Helsinki.

## Patient Consent

Written informed consent was obtained from all patients.

